# CARE-MVLM: Counterfactual Abstention and Region-Grounded Evidence in Mammography Vision-Language Models

**DOI:** 10.64898/2026.09.23.26363835

**Authors:** Bowen Qu, Weixin Liu, Matthew Murrow, Matthew Burger, Xingyi Guo, Mihir Sachin Vaidya, Susannah L. Rose, Murat Kantarcioglu, Bradley A. Malin, Zhijun Yin

## Abstract

Mammography interpretation requires tying each finding to supporting evidence and withholding judgment when that evidence is unavailable. Vision-language models are increasingly used for image-based medical question answering, yet existing benchmarks suggest that they struggle to jointly support accurate answers, evidence grounding, and appropriate abstention. We propose CARE-MVLM, built on Qwen2.5-VL-7B-Instruct, to address this gap through separate modules for answer prediction, answer-conditioned evidence generation, and answerability selection. On 7,358 triplets curated from CBIS-DDSM, MIAS, and VinDr-Mammo, CARE-MVLM substantially improves lesion-sensitive selective behavior over same VLM backbone baselines and exhibits markedly stronger lesion-sensitive abstention than GPT-5.6 and Gemini-3.5-flash while attaining the highest joint answer, grounding, and abstention reliability in the comparison.

## 1. INTRODUCTION

Breast cancer is the most frequently diagnosed cancer among women worldwide, and mammography remains the primary modality for screening and diagnostic assessment [1]. Reading a mammogram means more than naming a finding. Each reported abnormality must be tied to the supporting tissue region, and a reader who cannot see supporting evidence is expected to say so rather than guess. Vision-language models (VLMs) support image-based question answering [2], raising the prospect of mammography VLMs that behave similarly by answering questions about pathology and abnormalities, pointing to the lesion that explains each answer, and abstaining when evidence is lacking.

Existing methods address these requirements largely in isolation. Evidence-grounded medical VLMs tie predictions to image regions through phrase-conditioned localization [3], visual and textual explanations [4], and region-aware supervision [5]. Yet localizing a region does not establish that the region actually carries the predicted label. Selective prediction trains a model to withhold an answer [6, 7, 8], but abstention that is never tested against removal of relevant evidence cannot be distinguished from generic caution. A recent study replaces images with similar alternatives but reports accuracy drops of at most 6.5 points [9]. Since whole-image replacement changes both relevant and irrelevant content, response changes are difficult to attribute to specific evidence; even after tuning, 70% of shuffled images retain the same answer. This limitation is particularly relevant in mammography, where decisive evidence may be confined to a small lesion. Leading VLMs rarely pair correct answers with matching lesion evidence or abstain after that evidence is removed [10]. To our knowledge, no method jointly trains answer correctness, label-aware grounding, and evidence-conditioned abstention under a lesion-specific intervention.

In this paper, we propose CARE-MVLM, a lesion Counterfactual Abstention and Region-grounded Evidence Mammography VLM, which addresses these three requirements with separate modules (Fig 1). Each question is presented in an original view, a target-removal view in which the lesion is replaced with reconstructed tissue, and a random-removal view matched in size with the same editing operator. Trained and tested on triplets from CBIS-DDSM, MIAS, and VinDr-Mammo, CARE-MVLM achieves a 21.48-point target-versus-random abstention gap, compared with 8.24 for GPT-5.6 and 2.34 for Gemini-3.5-flash, while attaining the best answering-abstention balance. On the same backbone, grounded accuracy improves from 5.58% to 10.39% over grounded supervised fine-tuning.

**Fig. 1.**
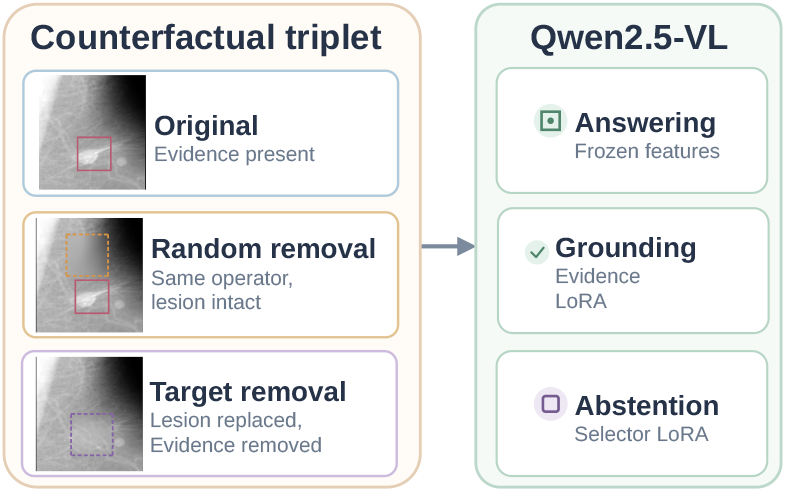
Overview of CARE-MVLM. It is trained using the original, random-removal, and target-removal views, with separate modules for answer prediction, evidence grounding, and abstention on a shared VLM backbone.

## 2. METHODS

## 2.1. Problem Formulation and Counterfactual Views

Given a mammogram x and a question *q*, CARE-MVLM predicts an answer *ŷ*with evidence boxes 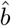, or abstains. Pathology selects one label from *Y*_P_ = *{Benign, Malignant}*, while Abnormality predicts a subset of the nine categories in *Y*_A_ such as *Mass* or *Calcification*. Counterfactual studies typically blank, blur, or swap out the queried region [11, 5, 9], creating a signature that remains detectable without reading anatomy. We follow [10] to generate target removal *x*^*T*^ by reconstructing each annotated lesion region from its surrounding non-lesion area [10], so no evidence or obvious masking signature remains in the reference answer. Random removal *x*^*R*^ applies the identical operator to lesion-free regions matched in count, width, and height, leaving the annotated lesions intact. Matching the editing operator and extent controls for these aspects of the intervention, although location- and tissue-dependent editing artifacts may still differ between the two views. Answer and evidence supervision uses the original view *x*^*O*^ and *x*^*R*^, while *x*^*T*^ supplies abstention targets and cross-view constraints, and thereby gives an operational definition of unanswerability for both training and evaluation.

### 2.2. Task-Specific Answer Prediction

Local encoders and task heads operate on frozen VLM features. Question-conditioned scores select up to *K*_*v*_ visual to-kens for weighted pooling. A multilayer perceptron (MLP) fuses the projected question representation *h*_*q*_ and pooled visual representation *h*_*v*_ as

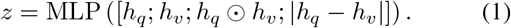

Pathology uses a softmax head, while Abnormality uses sigmoid outputs and a cardinality head that predicts how many labels to retain.

Here *L*_cls_ is weighted cross-entropy on *x*^*O*^ and *x*^*R*^, with focal modulation of the negative terms for the sparse Abnormality classes [12], and *L*_card_ a cross-entropy on the predicted label count. Three cross-view terms then shape the representation:

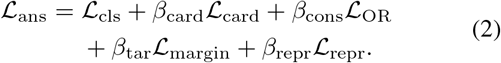

where *L*_OR_ penalizes prediction differences between *x*^*O*^ and *x*^*R*^, the margin loss separates ground-truth-label scores on *x*^*O*^ and *x*^*R*^ from those on *x*^*T*^ by *δ*_tar_, and the representation loss favors similarity between *x*^*O*^ and*x*^*R*^ over similarities between *x*^*O*^ and*x*^*T*^ and between *x*^*R*^ and*x*^*T*^, weighted by *β*_repr_.

### 2.3. Answer-Conditioned Evidence Generation

An independent low-rank adaptation (LoRA) adapter [13] for evidence generation models 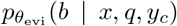, conditioning on ground-truth labels during training and on predicted labels at inference, so that localization is asked to explain a fixed answer rather than to compete with it. Coordinates are emitted as integers on a normalized grid, and the objective

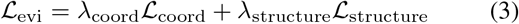

weights *L*_coord_ and *L*_structure_, token cross-entropies over box coordinates and over the remaining output tokens. Inference keeps only the geometrically valid boxes that match the predicted answer labels.

### 2.4. Counterfactual Answerability Selection

The selector combines a separate LoRA adapter and a prompt MLP with a question-conditioned local multiple instance learning (MIL) branch that pools up to *K*_*v*_ visual tokens:

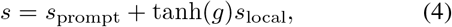

where *g* is a trainable gate. The selector receives no supervised answer tokens, ground-truth boxes, or view identities as inputs. It learns to accept *x*^*O*^ and *x*^*R*^ and to reject *x*^*T*^ through

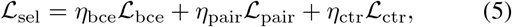

where *L*_bce_ is view-weighted binary cross-entropy on the acceptance target, *L*_pair_ ranks *x*^*O*^ and *x*^*R*^ above *x*^*T*^ by *δ*_pair_, and *L*_ctr_ contrasts representations with margin *δ*_ctr_. An offset *a* then controls acceptance relative to the threshold *τ*_acc_: the system returns 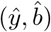 when *s*(*x, q*) + *a≥τ*_acc_ and abstains otherwise. The three modules are optimized in stages and frozen for inference. Base weights are never updated: the answer heads read backbone features directly, whereas the evidence and selector branches each insert their own adapter.

## 3. EXPERIMENTS

### 3.1. Data

The dataset draws mammograms, task labels, and lesion boxes from CBIS-DDSM, MIAS, and VinDr-Mammo [14, 15, 16], covering both digitized screen-film and full-field digital acquisitions; the nine-category abnormality label space follows the unified mapping of [17]. Each image–question instance is expanded into an original (*x*^*O*^), random-removal (*x*^*R*^), and target-removal (*x*^*T*^) triplet, giving 5,044 Train, 736 Eval, and 1,578 Test triplets. Both removal views use the same tissue-reconstruction operator, which extends the counterfactual construction of [10] to an operator-matched control. Test contains 622 binary Pathology questions and 956 potentially multi-label Abnormality questions.

### 3.2. Evaluation Metrics

Answer quality on *x*^*O*^ is reported as Pathology macro-F1 (**P-MF1**), Abnormality exact match (**A-EM**), and Abnormality macro-F1 (**A-MF1**), with abstentions receiving no correctness credit. Coverage (**Cov**.) is the proportion of valid non-abstaining answers and selective accuracy (**Sel-Acc**) is the accuracy among them; because a method can inflate Sel-Acc by answering nothing, we also report their product as the covered-and-correct rate (**CCR**), the fraction of all questions answered correctly. Grounded accuracy (**GA**) additionally requires every predicted box to match a label-compatible reference at an intersection over union (**IoU**) of 0.3 or higher, with all *x*^*O*^ as its denominator.

The counterfactual metrics compare behavior across the three views. The abstention specificity gap (**ASG**) is the target-removal abstention rate minus the random-removal abstention rate. Constant answering and constant abstention both yield zero ASG. With the editing operator and extent matched, a positive ASG indicates more frequent abstention after lesion removal than after non-target editing. **CF-H** is the harmonic mean of the target-removal abstention rate and the average correct-answer rate on *x*^*O*^*/x*^*R*^, and **GCF-H** replaces correct-answer rates with rates that additionally require strict localization. Joint reliability (**JR**) is the strictest criterion, requiring every requirement to hold within the same triplet:

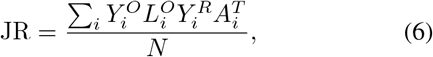

where *N* is the number of triplets,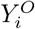 and 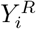 indicate correct non-abstaining answers on *x*^*O*^ and 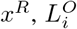 the same localization criterion used by GA, and 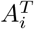 valid abstention on *x*^*T*^. Invalid outputs never count as valid abstentions. Paired ablation comparisons use pointwise 95% confidence intervals from 2,000 bootstrap resamples of the Eval benchmark groups, which keep grouped questions and views together.

### 3.3. Models and Training

All local methods use Qwen2.5-VL-7B-Instruct [18] as a backbone, ranging from fine-tuning to counterfactual supervision. B0 applies zero-shot learning. B1 supervises answers on the original views only. B2 adds evidence-box supervision, so it is grounded but never sees a removal view. B3 additionally supervises abstention on target-removal views, but does so inside a single generator with no separate selector, which is the natural way to add abstention without the proposed decomposition. CARE-MVLM w/o selector disables selection while keeping identical answer and evidence modules, isolating what the selector contributes. GPT-5.6 and Gemini-3.5-flash are evaluated on the same triplets under the same evaluation protocol, at a temperature of 0. LoRA uses rank 16, and each batch contains 16 triplets. The answer heads are trained for five epochs at a learning rate of `3e-4` and the final evidence refinement for two epochs at `1e-5,` with seed 42. Two hyperparameters are tuned on Train-dev: the coordinate weight *λ*_coord_ *∈* {0.2, 0.5, 0.8}, selected by generator GA with mean matched IoU breaking ties, and the acceptance offset *a∈* {− ln 4, *−*ln 2, 0, ln 2, ln 4}, selected by JR with CF-H breaking ties. Test uses the settings fixed before evaluation, *λ*_coord_ = 0.8 and *a* = ln 2.

### 3.4. Results

Table 1 summarizes model performance across four evaluation categories. Answer accuracy and evidence-grounded behavior clearly diverge. B1 and B2 achieve 56.08% and 55.64% Sel-Acc at full coverage, yet neither abstains after lesion removal, leaving all four counterfactual metrics at zero. B2 differs from B1 only by adding box supervision, which increases GA from 0.00% to 5.58% without improving counterfactual behavior, indicating that adding localization supervision does not improve measured counterfactual abstention. B3 exhibits severe over-abstention. Its 76.00% Sel-Acc is the highest, but it answers only 1.58% of questions, yielding a CCR of 1.20% compared with 56.08% for B1. This result highlights the need to report coverage alongside selective accuracy and motivates evaluating a separate selector.

**Table 1.**
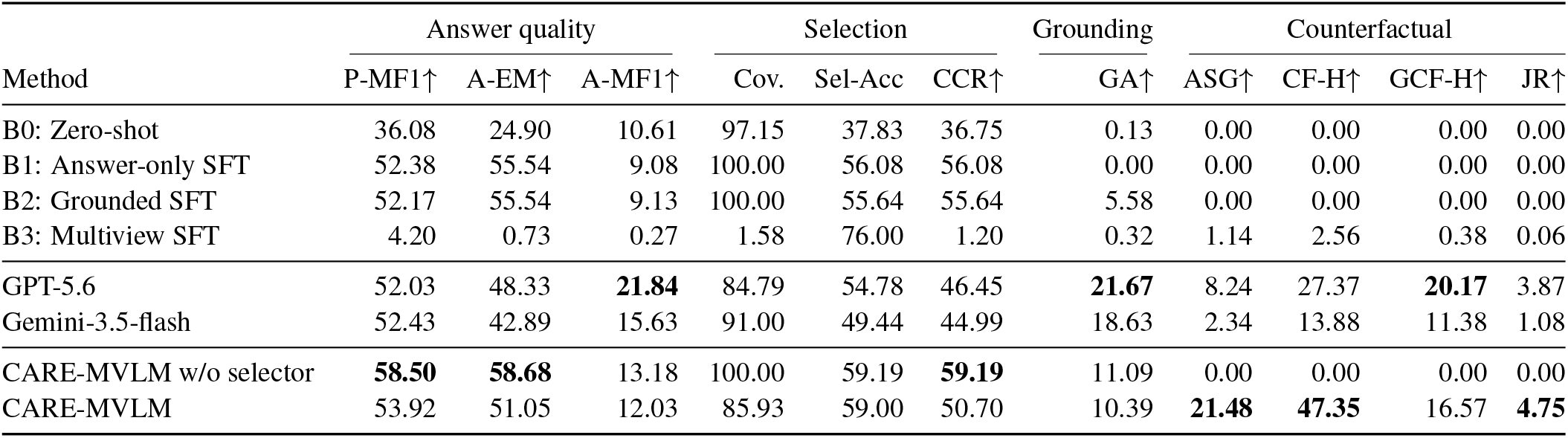
Results on the test set. Answer, selection, and grounding metrics use the original views, while the counterfactual metrics use complete triplets. All values are scaled by 100; ASG is the difference between two abstention rates. GA uses an IoU threshold of 0.3. Coverage and Sel-Acc carry no arrow because they are operating points that lead to CCR, which credits a method only for questions it both answers and answers correctly. Bold indicates the best value for each metric marked with *↑*.

CARE-MVLM reaches an ASG of 21.48 percentage points compared with at most 1.14 for the same-backbone baselines, and raises GA from 5.58% for B2 to 10.39%. The positive ASG is consistent with greater sensitivity to lesion removal than to matched non-target editing. Compared with CARE-MVLM w/o selector, enabling selection raises target-removal abstention from 0.00% to 47.97%, while original-view coverage decreases from 100.00% to 85.93% and CCR from 59.19% to 50.70%. GA decreases from 11.09% to 10.39%, and Sel-Acc remains similar at 59.00% versus 59.19%. Thus, the selector improves counterfactual abstention but does not improve original-view selective accuracy, consistent with its objective of predicting intervention-defined answerability rather than answer correctness. JR reaches 4.75%, compared with at most 0.06% for the same-backbone baselines.

Compared with GPT-5.6 and Gemini-3.5-flash, CAREMVLM achieves higher Sel-Acc and CCR, at 59.00% and 50.70%, respectively. It also achieves a larger ASG, at 21.48 percentage points versus 8.24 and 2.34, and a higher CF-H, at 47.35 versus 27.37 and 13.88. Its JR point estimate is the highest at 4.75%, compared with 3.87% and 1.08%. However, its GA is lower at 10.39%, compared with 21.67% and 18.63%. Its GCF-H is 16.57, which is lower than GPT-5.6 (20.17) but higher than Gemini-3.5-flash (11.38). Among original views that are both correctly answered and grounded, correct answering under random removal and abstention under target removal jointly hold for approximately 46% of cases, against 18% for GPT-5.6 and 6% for Gemini-3.5-flash; this ratio is obtained as JR divided by GA.

Conditioning on original views that are correctly answered and strictly grounded helps distinguish sensitivity to lesion removal from generic sensitivity to editing. Among these cases, all systems change answers far more often after target removal than after random removal: 62.20% against 12.20% for CARE-MVLM, 71.35% against 16.37% for GPT-5.6, and 71.77% against 13.61% for Gemini-3.5-flash. However, 88% of the CARE-MVLM changes after target removal are abstentions, compared to 28% for GPT-5.6 and 9% for Gemini-3.5-flash, which mainly switch to a different label. CARE-MVLM also rarely localizes to the edited region on random-removal views, with only 1.58% of predicted boxes lying at least half within the edited region, compared with 11.79% for GPT-5.6 and 13.24% for Gemini-3.5-flash.

In addition, we asked a radiologist to review 50 random triplets. Target removal completely removed the lesion in 92% of cases, and random removal preserved diagnostically relevant evidence in 94%. No interpretation-changing artifact was identified in 76%; edit-related asymmetry occurred in 10%, while the remaining 14% showed other editing artifacts, including nipple removal and altered tissue appearance. No new mass, calcification, architectural distortion, or malignant-appearing lesion is reported. These findings support the validity of the counterfactual construction while highlighting occasional limitations of synthetic editing.

### 3.5. Ablation Study

The ablations focus on two design choices: conditioning evidence generation on the answer, and the pairwise and contrastive terms in the selector objective. Each ablation is compared against its own matched reference run, sharing initialization, frozen components, and update count within that comparison but not across the two, which is why the two reference values of GCF-H differ. Removing answer conditioning reduces GCF-H from 17.00 to 14.90, a paired difference of *−*2.10 points with a 95% confidence interval of [*−*3.74, *−*0.49], while CF-H is unchanged because answers and selector decisions are fixed; the contribution is therefore specifically to grounding. Removing the pairwise and contrastive terms together reduces CF-H from 48.41 to 45.26 and GCF-H from 17.37 to 16.76, paired differences of *−*3.15 points ([*−*5.32, *−*1.11]) and *−*0.61 points ([*−*1.17, *−*0.06]), so the counterfactual balance depends on the ranking and contrastive structure of the selector rather than on binary abstention supervision alone.

## 4. CONCLUSION

CARE-MVLM jointly trains answering, evidence grounding, and answerability selection on lesion-counterfactual triplets. It achieves the strongest lesion-sensitive abstention and highest joint reliability across same-backbone baselines and proprietary VLMs, showing the value of explicitly training evidence-dependent behavior. Joint reliability remains low in absolute terms, with evidence grounding a key target for improvement. Future work should evaluate additional VLMs and more natural evidence-absence settings. Overall, CARE-MVLM provides a practical framework for jointly training and evaluating evidence-grounded answering and appropriate abstention in medical VLMs.

## Data Availability

The data analyzed in this study were obtained through MammoVQA and originate from CBIS-DDSM, MIAS, and VinDr-Mammo. All data were publicly available before the initiation of the study. Access information and citations for the datasets are provided in the manuscript and through the links below. No newly collected or identifiable patient data were used.

https://github.com/PiggyJerry/MammoVQA

https://www.cancerimagingarchive.net/collection/cbis-ddsm/

https://www.repository.cam.ac.uk/items/b6a97f0c-3b9b-40ad-8f18-3d121eef1459

## Notes

### Competing Interest Statement

The authors have declared no competing interest.

### Author Declarations

This study used only existing, de-identified human data that were openly available to the public before the initiation of the study. The analyzed MammoVQA records and associated mammographic images originate from CBIS-DDSM, MIAS, and VinDr-Mammo and were obtained from publicly accessible repositories without application, screening, registration, or individual request. No new patient data were collected, and no identifiable patient information was accessed. Links to the source data are provided below.

### Summary of Updates

Minor wording and typographical errors were corrected.

